# A Multicentre Randomised Controlled Trial Assessing the Efficacy of Antimicrobial Prophylaxis for Extracorporeal Shock Wave Lithotripsy in Reducing Urinary Tract Infection (APPEAL): Statistical Analysis Plan and Methodology

**DOI:** 10.1101/2025.03.09.25323609

**Authors:** Philippe D. Violette, Farhad Tondro Anamag, Sameer Parpia, Sara V. Tornberg, Arto Mikkola, Sakineh Hajebrahimi, Saana Horstia, Borna Tadayon Najafabadi, Jani Ruotsalainen, Pramila Gaudel, APPEAL Trial Investigators, Le Mai Tu, Thomas Tailly, Farzin Soleimanzadeh, Hanieh Salehi Pourmehr, Mari Saalasti, Patrick O. Richard, Hassan Razvi, Negar Pourjamal, Stephen E. Pautler, Sanna Myrskysalo, Andrei O. Morozov, Mohsen Mohammadrahimi, Murilo de Almeida Luz, Samuel Lagabrielle, Pauliina Kuutti, Tuomas P. Kilpeläinen, Petrus Järvinen, Alireza Farshi Haghro, Salam A. Hussain, Agus Rizal A.H. Hamid, Dmitry Gorelov, Nariman Gadzhiev, John Denstedt, Kathrin Bausch, Raed A. Azhar, Moza Al Hail, Nourieh D. Akbari, Mohamed A. AbdElAziz, Mohamed Abdelkareem, Gordon H. Guyatt, Kari A. O. Tikkinen

**Affiliations:** Department of Health Research Methods, Evidence and Impact, McMaster University, Hamilton, ON, Canada; Division of Urology, Department of Surgery, McMaster University, Hamilton, ON, Canada; Department of Surgery, Woodstock Hospital, Woodstock, ON, Canada; Department of Urology, Tabriz University of Medical Sciences, Tabriz, Iran; Research Center for Evidence-Based Medicine, Faculty of Medicine, Tabriz University of Medical Sciences, Tabriz, Iran; Faculty of Medicine, University of Helsinki, Helsinki, Finland; Department of Oncology, McMaster University, Hamilton, ON, Canada; Department of Urology, University of Helsinki and Helsinki University Hospital, Helsinki, Finland; Division of Urology, Department of Surgery, University of Toronto, Toronto, ON, Canada; Division of Urology, Department of Surgery, Centre Hospitalier Universitaire de Sherbrooke, Sherbrooke, QC, Canada; Department of Urology, University Hospital Ghent, Ghent, Belgium; Clinical Trials Unit, HUS Pharmacy, Helsinki University Hospital, Helsinki, Finland; Division of Urology, Schulich School of Medicine and Dentistry, Western University, London ON, Canada; Institute for Urology and Reproductive Health, Sechenov University, Moscow, Russia; Department of Urology, Rede D’Or São Luiz, Sao Paolo, Brazil; Urology Section, Surgery Department, Hazm Mebaireek General Hospital, Hamad Medical Corporation, Doha, Qatar; Department of Urology, Faculty of Medicine, Universitas Indonesia, Cipto Mangunkusumo Hospital, Jakarta, Indonesia; Department of Endourology, Pavlov First St. Petersburg State Medical University, St. Petersburg, Russia; Department of Urology, Saint Petersburg State University Hospital, St. Petersburg, Russia; Department of Urology, University Hospital Basel, Basel, Switzerland; Department of Urology, Faculty of Medicine, King Abdulaziz University, Jeddah, Saudi Arabia; Department of Medicine, McMaster University, Hamilton, ON, Canada; Department of Surgery, South Karelian Central Hospital, Lappeenranta, Finland; Department of Surgery, Päijät Häme Central Hospital, Lahti, Finland

**Author notes:** **Correspondence:** Kari Tikkinen. Faculty of Medicine, University of Helsinki, Biomedicum 2B, P.O. Box 13, Tukholmankatu 8 B, 00290 Helsinki, Finland. **Collaborator authors** (APPEAL Trial Investigators): Alex L. E. Halme, Niko K. Nordlund, Paula Saari, Linda Nott, Elsie Morneau, Behzad Lotfi, Mohsen Amjadi, Kamaleddin Hassanzadeh Nokashti, Nima Naghdi, Etham Jahantabi, Ehsan Sepehran, Fatimeh Kazemi Rashed, Zeenah Abdulmajid, Khalid Al-Rumaihi, Helge Seifert, Maeve Dreher, Aiman Al-Solumany, Espie Quiroz, Mustafa Taeyb, Wissam Kamal, Ahmad Bugis, Alexandra Mischenko, Dmitry Lebedev, Vadim Rudenko, Nataliya Antsupova, Dmitry Enikeev, Felipe A Loprete, Carlos A Guedes Negrão, Yasmina Zahra Syadza, Widi Atmoko, Bayu Hernawan, Ervita Mediana, Annisa Vigilanty Pratiwi, Rustom P Manecksha, Lorraine Scanlon, Andreia Bilé Silva.

**Keywords:** antibiotic prophylaxis, clinical trials, lithotripsy, randomized controlled trial, statistical analysis, urinary tract infections, urolithiasis

## Abstract

**Background:** Urinary tract infection (UTI) is a recognized complication of shock wave lithotripsy (SWL) for urolithiasis. Evidence guiding antibiotic prophylaxis remains of low certainty, contributing to substantial practice variation and conflicting guidelines. A well-powered, blinded randomized trial is essential to provide trustworthy evidence for clinical practice.

**Study Design:** An international, multicentre, randomized controlled trial (the APPEAL trial) assessing the benefits and harms of a single dose of ciprofloxacin versus placebo before SWL in reducing post-procedure UTI. Patients, healthcare providers, data collectors, outcome adjudicators and statisticians blinded to treatment assignment.

**Endpoints:** Appeal’s primary outcome is bacteriuria or symptomatic UTI (symptomatic UTI defined as symptomatic cystitis, pyelonephritis, or urosepsis) within approximately 7-14 days post-SWL. Other outcomes include pyelonephritis or urosepsis, and serious adverse events.

**Patients and Methods:** Over 1,500 patients from high- and middle-income countries undergoing SWL for nephrolithiasis or ureterolithiasis. Exclusion criteria include a positive pre-SWL urine analysis for nitrites or urine culture, ongoing or planned antibiotic use, suspected struvite stones, urinary catheters or diversion, or a history of urosepsis.

Imminent report of APPEAL will provide high-quality evidence on the role of antibiotic prophylaxis in SWL and identify subgroups that may benefit most.

**Trial Registration:** ClinicalTrial.gov identifier (ID): NCT03692715.

## Background

Urolithiasis is a common urological condition that is associated with considerable morbidity and significant economic burden [1,2]. Shock wave lithotripsy (SWL), a less invasive treatment than ureteroscopy, percutaneous nephrolithotomy, and other surgical treatments for renal and ureteral calculi, carries a risk of complications, notably infection [3–8]. In patients undergoing SWL, potential risk factors for urinary tract infections (UTI) or bacteriuria include having ureteral stent, large stone size, high stone density, kidney/upper ureteral (vs distal ureteral) stones, older age, female (vs male) sex, and diabetes mellitus [5–8].

### Clinical practice

A 2016 survey comparing practice patterns between Canadian (n=94) and American (n=187) urologists found that routine pre-SWL antibiotic use was much more common in the United States (78%) than in Canada (2%) [9]. In a UK retrospective study (data collection starting September 2013) most patients did not receive antibiotic prophylaxis pre-SWL. However, clinicians from two (out of seven) centers routinely administered 3-5 day post-treatment prophylaxis [10].

As there is paucity of studies on practice patterns, we surveyed each of the 12 trial centers in 9 countries before they initiated recruitment. Investigators in 3 (25%) centers reported typically not prescribing antibiotic prophylaxis for patients. In 4 (33%) centers, investigators prescribed prophylaxis only for patients with a ureteral stent, while in 5 (42%) centers, investigators typically prescribed prophylaxis regardless of the presence of a stent. Appendix 1 provides details on our survey methods and results. The practice variation identified by previous studies [9,10], along with findings from our own survey, underscore the need for conducting our randomized trial.

### Guidelines and evidence

The European Association of Urology (EAU) Guidelines on Urological Infections gives a strong recommendation as “*Do not use antibiotic prophylaxis to reduce the rate of symptomatic urinary infection following extracorporeal shockwave lithotripsy*” [11]. The EAU Guidelines on Urolithiasis states that “*No standard antibiotic prophylaxis before SWL is recommended. However, prophylaxis is recommended in the case of internal stent placement ahead of anticipated treatments and in the presence of increased bacterial burden*” [12]. The Canadian Urological Association (CUA) gives a moderate recommendation that states: “*Pre-procedural antibiotics do not significantly reduce the risk of UTI and fever in patients undergoing ESWL, but should be considered in patients at high risk of infectious complication*s” [13]. The American Urological Association (AUA) 2020 Best Practice Statement states that “*non-invasive procedures such as shock wave lithotripsy do not require antimicrobial prophylaxis if the pre-procedural urine microscopy is negative for infection*”, without making any specific recommendations for any potential higher risk groups [14].

The EAU urolithiasis and urological infections guidelines [11,12] base their recommendations primarily on two systematic reviews and meta-analyses [13,15], one published in 2012 (9 RCTs with 1364 patients included in the meta-analysis on UTI) and the other in 2015 (7 RCTs with 820 patients included in the meta-analysis on UTI). Although these meta-analyses concluded that antibiotic prophylaxis did not reduce UTI or fever, the 2012 meta-analysis that reported 41 events for UTI, demonstrated a relative risk reduction of 46% (RR 0.54, 95% CI 0.29-1.01), and the 2015 meta-analysis that reported 42 events for fever, demonstrated a relative risk reduction of 74% (RR 0.26, 95% CI: 0.06-1.1). This low certainty evidence (due to imprecision and other factors) may actually suggest benefit of antibiotic prophylaxis in reducing UTI or fever. The AUA guideline classified its recommendation as clinical principle without providing supporting evidence [16].

The most recent systematic review examining this topic [17], published in 2021, included a meta-analysis of 1,482 patients who experienced 43 symptomatic UTI events. Authors rated the evidence certainty as very low and reported a RR of 0.55 (95% CI 0.22–1.36). The authors concluded that antibiotic prophylaxis is not necessary for patients with sterile urine prior to ESWL [17]. However, due to the very low certainty of evidence, high risk of bias, and a low number of events resulting in a wide confidence interval, whether antibiotic prophylaxis confers important benefits remains uncertain. Resolving this uncertainty and establishing clear evidence-based practices that strike a balance between minimizing antibiotic prophylaxis use and preserving antimicrobial effectiveness without increasing postoperative complications requires a large, randomized trial. In this context, the APPEAL trial will compare the efficacy of single-dose ciprofloxacin antibiotic prophylaxis with placebo in preventing postoperative bacteriuria and urinary tract infections (cystitis, pyelonephritis, and urosepsis) post SWL for urolithiasis.

## Study Design

### Design and management

The APPEAL trial (Table 1) is a phase IV placebo-controlled, parallel-arm, international, multicentre, randomized controlled trial comparing preoperative single-dose ciprofloxacin to placebo in patients undergoing SWL for urolithiasis. A clinical trial update article provides information on study progress [18]. The present manuscript focuses on the methods of the APPEAL trial and provides the statistical analysis plan.

**Table 1.** Administrative information.

|  |  |
| --- | --- |
| **Official Title** | A Multicentre Randomized Controlled Trial Assessing the Efficacy of Antimicrobial Prophylaxis for Extracorporeal Shock Wave Lithotripsy on Reducing Urinary Tract Infection (APPEAL) |
| **Brief Title** | Efficacy of Antimicrobial Prophylaxis before Shock Wave Lithotripsy (APPEAL) |
| **ClinicalTrials.gov Registration** | NCT03692715; https://www.clinicaltrials.gov/study/NCT03692715<br>(registered 02 October 2018) |

The Clinical Urology and Epidemiology (CLUE) Working Group at the University of Helsinki and Helsinki University Hospital leads and coordinates the APPEAL trial (as a coordinating centre). The CLUE Working Group is responsible for overseeing the trial, including study database and data consistency, statistical expertise and analysis, and the coordination of participating centres.

Grants from the Competitive Research Funding of the Helsinki and Uusimaa Hospital District, Sigrid Jusélius Foundation, Research Council of Finland, and Vyborg Tuberculosis Foundation support the APPEAL trial. The sponsors have no role in the study design or the manuscript preparation, review, or approval. The coordinating centre and the APPEAL trial Steering Committee are solely responsible for the design, data analysis, manuscript writing, and decision to submit for publication of this trial. The APPEAL trial is conducted in compliance with the study protocol, the Declaration of Helsinki, the International Council for Harmonization - Good Clinical Practice (ICH-GCP), and registered with ClinicalTrials.gov (Table 1). At each site, before the start of the recruitment, the institutional ethics boards approved the protocol that follows national legal and regulatory requirements. Appendix 2 provides information on decisions/approvals of the local institutional review boards.

### Recruitment

APPEAL is a multinational trial with centres representing four WHO regions (Americas, South-East Asia, Europe, and Eastern Mediterranean) (Figure 1). The 12 participating centres are from 9 countries including Finland (Helsinki University Hospital, Helsinki; as the coordinating centre), Canada (Western University Hospital, London, Ontario; and Centre Hospitalier Universitaire de Sherbrooke, Sherbrooke, Quebec), Iran (Tabriz University of Medical Sciences, Tabriz), Qatar (Hamad Medical Corporation, Doha), Russia (State Pavlov Medical University, Saint Petersburg; Aleksandrovskaya Hospital, Saint Petersburg; and Sechenov First Moscow State Medical University, Moscow), Switzerland (University Hospital Basel, Basel), Saudi Arabia (King Abdulaziz University Hospital, Jeddah), Brazil (Hospital E Maternidade São Luis, Sao Paolo), and Indonesia (Universitas Indonesia, Jakarta). These centres represent urologic departments with vast experience in SWL treatment of kidney and ureteral stones. Before starting the trial, eligible trial sites provided reassurance that ciprofloxacin resistance is not substantial in their patient population.

**Figure 1.**
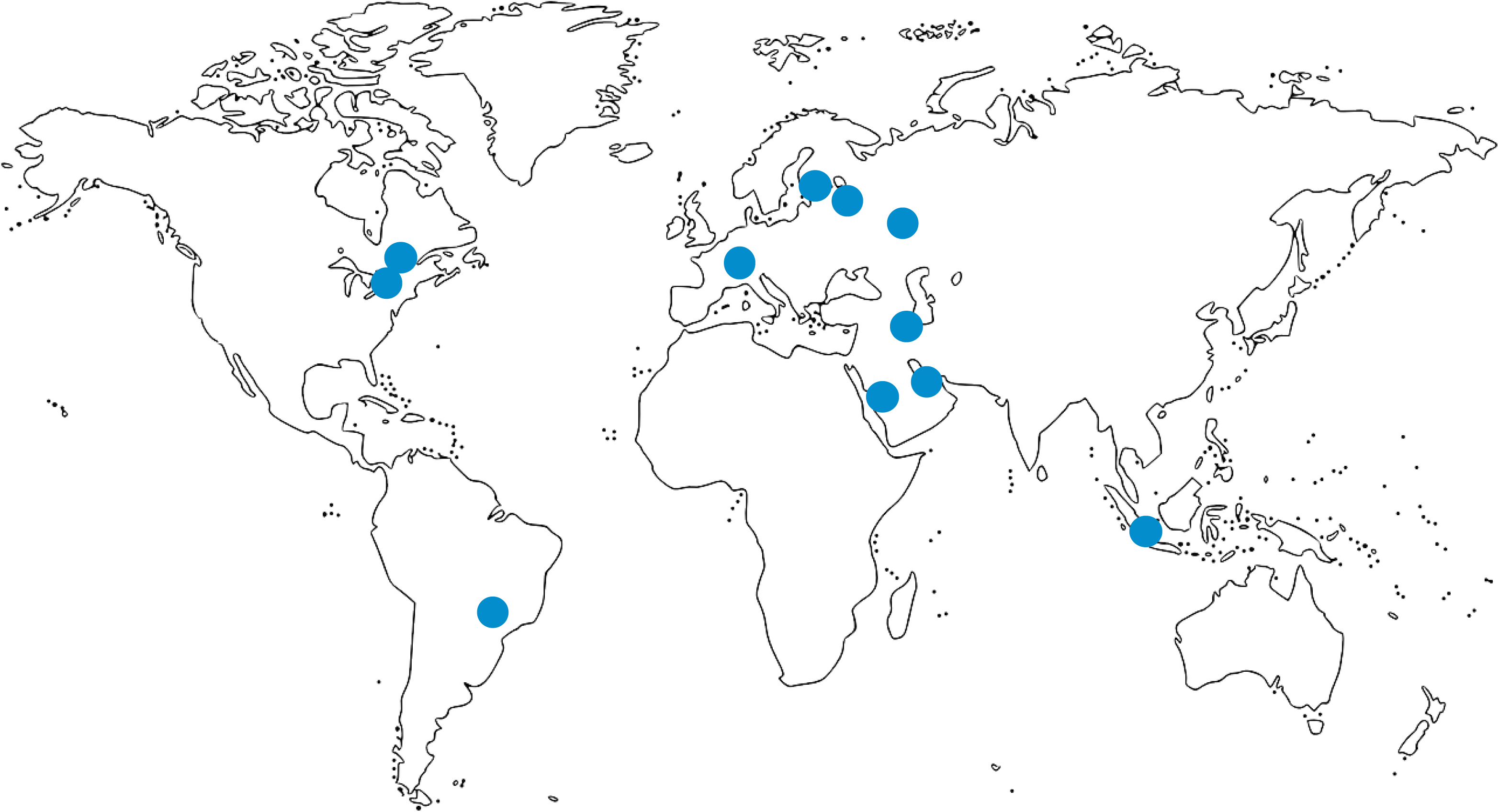
Recruiting centers of the APPEAL trial.

### Randomization and blinding

On day of the operation, randomized patients receive either a single dose of antibiotic prophylaxis with ciprofloxacin or a single dose of placebo pre-SWL (1:1 ratio with stratification by study site). Block randomization using blocks of 10, carried out by a central randomization organization (www.randomize.net, Ottawa, Canada) using a computerized algorithm is accessible via a standard browser on any internet-connected device. Patients are assigned a randomization number that corresponds to their treatment arm. The study treatment pack is assigned to the patient. Un-blinding will occur only after the completion of the statistical analysis. Patients, healthcare providers, data collectors, and outcome adjudicators are blind to active-ciprofloxacin or placebo-ciprofloxacin treatment assignment.

### Endpoints

We defined the primary outcome as a composite of positive urine culture, symptomatic cystitis, pyelonephritis or urosepsis at approximately 7-14 days (Table 2). We defined secondary outcome as any symptomatic urinary tract infection (composite of symptomatic cystitis, pyelonephritis or urosepsis), and tertiary outcomes as all individual components of the primary composite outcome as well as serious adverse events (SAEs). Table 2 presents a detailed description of the components and definitions of the primary, secondary, and tertiary outcomes.

**Table 2.** Outcomes and their definitions.

| Variable | Description |
| --- | --- |
| **Outcome definitions** |  |
| Bacteriuria | A significant number of bacteria (typically of a single uropathogen $\geq 10^5$ colony forming units/ml) in a urine culture |
| Symptomatic cystitis | Significant bacteriuria accompanied by the new onset of urinary frequency, urgency, and/or pain or burning<sup>a</sup> |
| Pyelonephritis or urosepsis | Positive post-SWL urine culture with fever ($>38.5^\circ\text{C}$ or $>101.1^\circ\text{F}$) likely due to urinary tract infection<sup>b</sup> |
| Serious adverse events | Any adverse event that results in hospitalization, is life-threatening, causes significant disability or incapacity, or requires intervention to prevent permanent impairment. |
| **Outcomes** |  |
| *Primary outcome* | Bacteriuria OR symptomatic cystitis OR pyelonephritis or urosepsis at approximately 7-14 days |
| *Secondary outcome* | Symptomatic cystitis OR pyelonephritis or urosepsis at approximately 7-14 days |
| *Tertiary outcomes* | Individual components of the primary outcome (bacteriuria, symptomatic cystitis, and pyelonephritis or urosepsis) |
|  | Serious adverse events |
| <sup>a</sup> These symptoms are assessed pre-SWL and at 7-14 days post-SWL using the following questions:<br>Urinary frequency: "How often have you had to urinate again less than two hours after finishing?"<br>Urinary urgency: "How difficult have you found it to postpone urination?"<br>Pain/Burning: "How often have you experienced burning or discomfort with urination?"<br>All questions use a six-point scale (0 = Not at all, to 5 = Almost always). New onset is defined as a score at least 1-point higher post-SWL compared to pre-SWL (Appendix 3). |  |
| <sup>b</sup> If a patient receives antibiotics post-SWL for suspicion of pyelonephritis or urosepsis (meaning that the patient has fever) without providing post-SWL urine culture, and without signs or symptoms of any other infection, such as upper respiratory tract infection, we consider the patient as having pyelonephritis or urosepsis. If a patient receives antibiotics post-SWL without providing post-SWL urine culture and has fever and symptoms of other infection than urinary tract infection, such as upper respiratory tract infection, we consider the patient lost to follow up. If a patient receives antibiotics post-SWL without providing post-SWL urine culture and without fever, we consider the patient lost to follow up. |  |

## Eligibility Criteria

Eligible patients are aged 18 years or older, have urolithiasis (i.e., nephrolithiasis, ureterolithiasis, or both), and undergo SWL. Table 3 lists the complete inclusion and exclusion criteria. All study participants provide written informed consent before randomization.

**Table 3.**
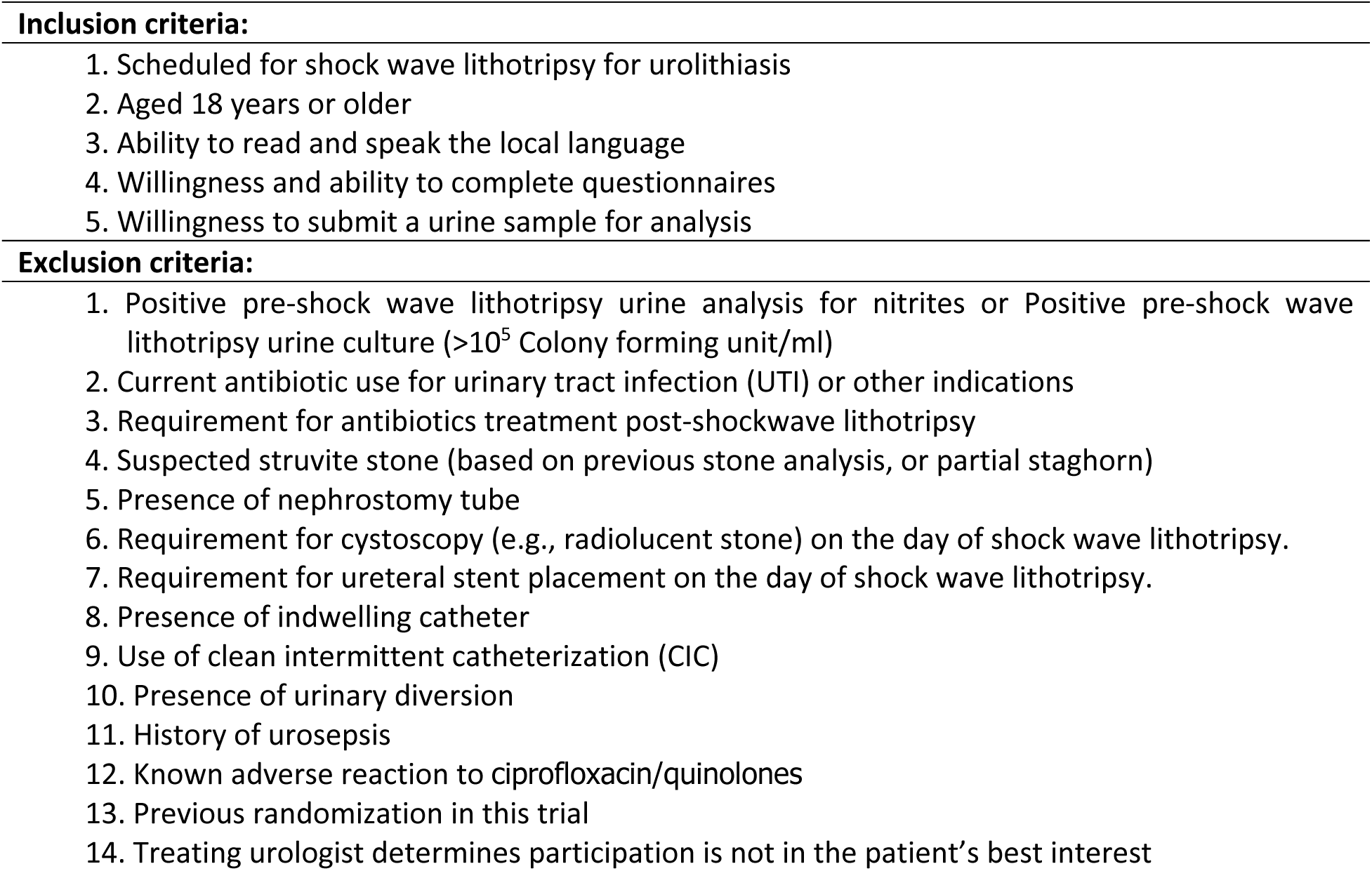
Eligibility (inclusion and exclusion) criteria.

## Methods

Pre-SWL patients are randomized to either antibiotic prophylaxis with ciprofloxacin or a single dose of placebo. Patients randomized to the ciprofloxacin group are administered a single dose either orally 500 mg (Finland, Switzerland) or intravenously 400 mg (Canada, Iran, Qatar, Russia, Saudi Arabia, Brazil, and Indonesia). Depending on the administration route chosen, the placebo group receives either a single tablet identical to ciprofloxacin or intravenous saline without any added active pharmaceutical. Patients receive oral intervention approximately one hour prior to the beginning of the SWL and the intravenous intervention immediately prior to SWL.

We exclude patients with known allergy for ciprofloxacin or quinolones (Table 3). We discuss risks and benefits of receiving either the trial intervention or placebo with patients and inform them of the low risk of allergic reaction, colonization with antibiotic resistant organisms, and the very low risk of developing of C. Difficile related diarrhoea, or tendon injury.

### Sample Size Calculation

To estimate the expected incidence of the primary outcome, we used the incidence of bacteriuria reported in the AUA Best Practice Statement on antibiotic prophylaxis: patients undergoing SWL in the placebo arm of trials assessing antibiotic prophylaxis had a 5.7% rate of bacteriuria as compared to 2.1% among those who received prophylaxis [19]. This represents an absolute difference of 3.6% (a 60% relative risk reduction). Therefore, to achieve a power of 90% (1 – β = 0.9) and to find an absolute difference of 3.6% in the occurrence of bacteriuria, with a significance level of α = 5% two-sided, we need to recruit 661 patients in each arm for a total of 1,322 patients.

Accounting for 10% loss to follow-up, the total required will be 1,454, or approximately 1,500 patients.

### Methods of Data Collection

Prior to SWL, we collect data through a pre-procedural study questionnaire, a modified IPSS questionnaire (Appendix 3), and obtain a urine sample (Table 4). A pre-procedural questionnaire, recorded at the time of SWL at the participating centre addresses clinical issues such as the presence or absence of a double J stent, stone characteristics, and renal insufficiency. At approximately 7-14 days post-SWL, the participants complete a modified IPSS questionnaire and provide urine for culture. Data acquisition and the occurrence of outcomes are monitored continuously through scheduled audits (Table 4).

**Table 4.**
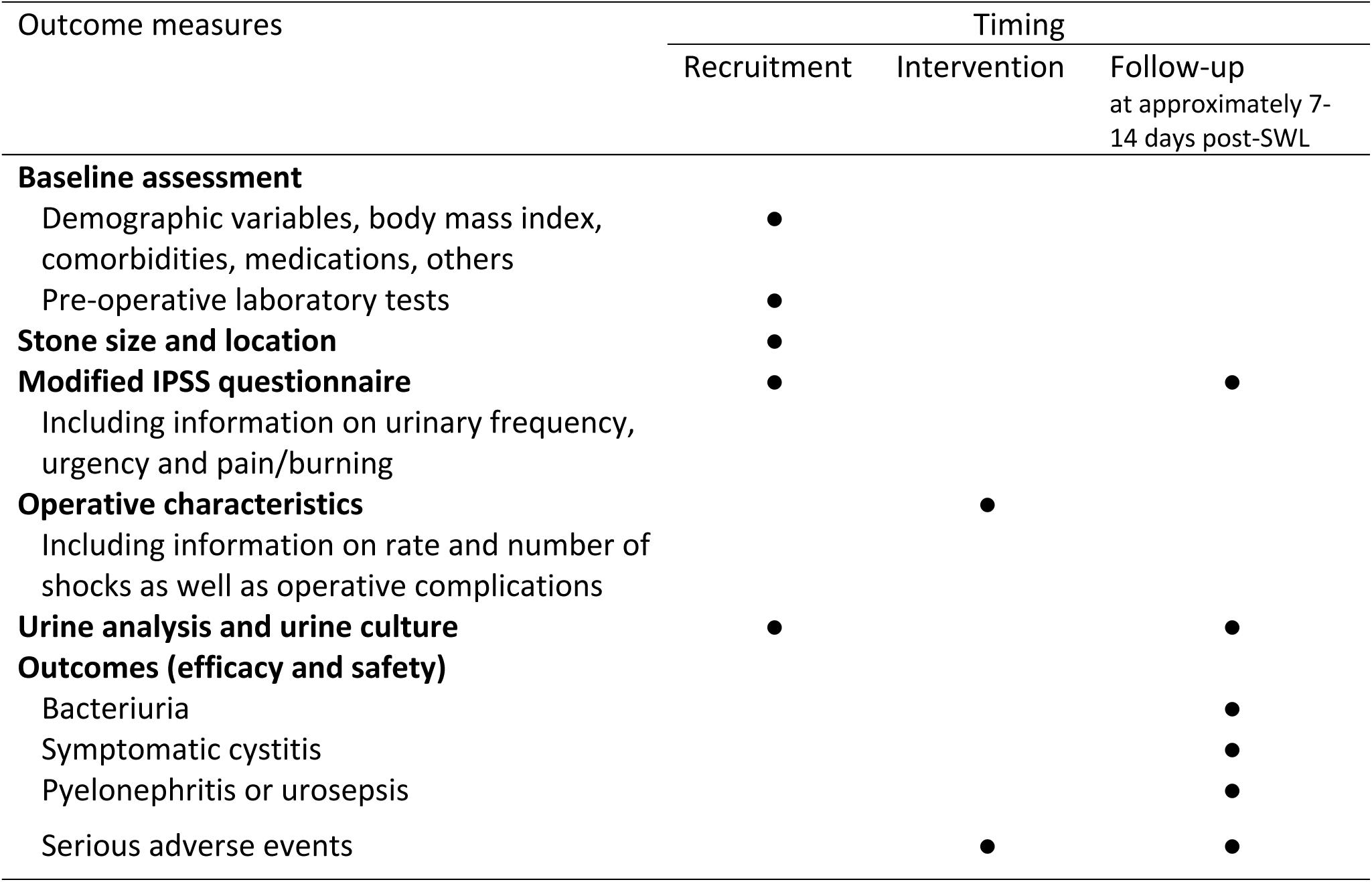
Schedule for data collection and outcome assessment.

### Analysis Plan

We will perform all outcome analyses using the intention-to-treat (ITT) principle, that is, we will analyse patients in the treatment group into which they were allocated (ciprofloxacin versus placebo), regardless of the intervention received.

#### Main Analyses

We will report baseline characteristics using medians (with interquartile ranges, IQR), mean values (with standard deviations), or frequencies (percentages). We will present all binary outcomes as crude numbers and proportions of participants in each group with the event (Table 5). We will conduct the primary analysis using a complete case approach, with participants having missing data excluded from the analysis. For the primary and secondary outcomes, we will use multivariable robust Poisson regression models controlling for study site, and presence (vs. absence) of ureteral stent. We will also conduct the same analysis for tertiary outcomes, provided there is an adequate number of events. The robust Poisson regression model uses the classical sandwich estimator under the generalized estimation equation (GEE) framework to provide accurate standard errors for the elements [20,21]. We will report results as relative risk reduction and absolute risk reduction with corresponding two-sided 95% confidence intervals (CIs) and associated p values. We will infer statistical significance if the computed 2-sided *p*-value is <0.05. We will conduct blinded interpretation of study results: i) we will first conduct blinded review of primary outcome data to develop two interpretations, one assuming A as the experimental treatment (ciprofloxacin) and another as the control (placebo). After finalizing and signing decisions, we will break the randomization, and the correct interpretation is selected. We will perform analysis using Stata version 18 (StataCorp LLC, College Station, TX, USA, 2023) and R version 4.4.2 (R Core Team, 2024; R Foundation for Statistical Computing, Vienna, Austria).

**Table 5.**
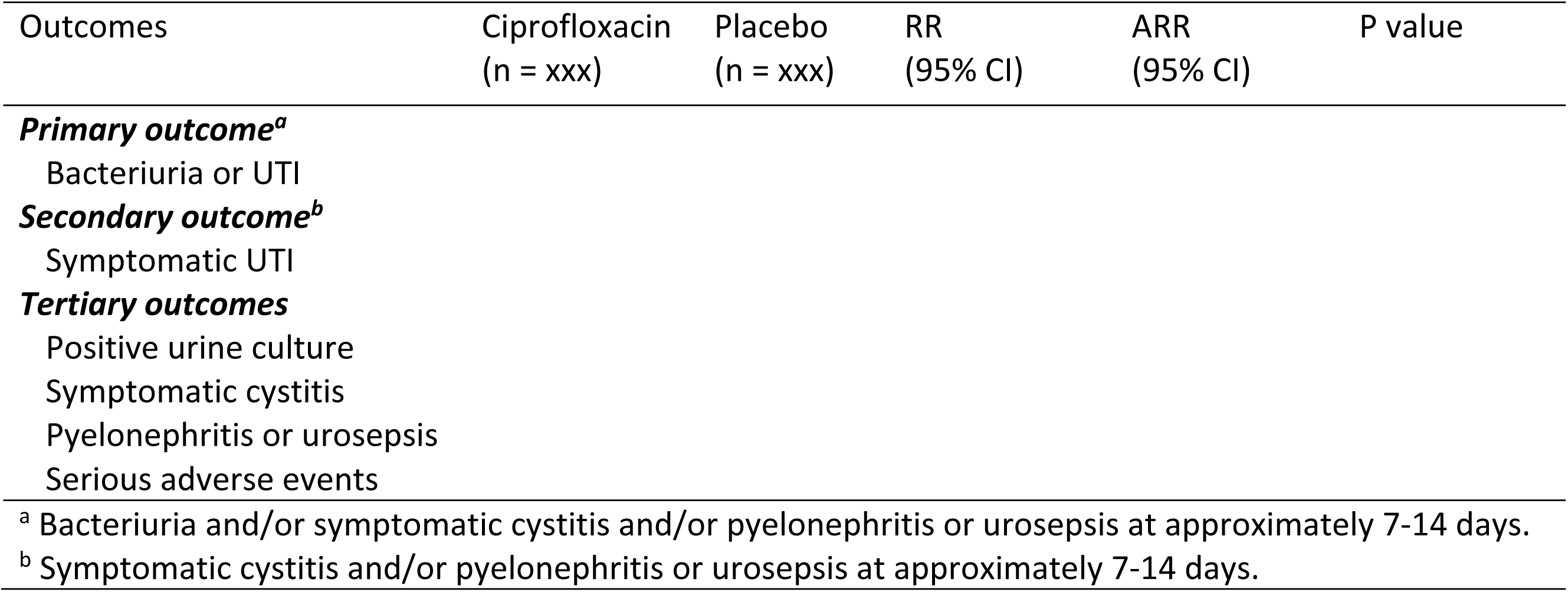
Study outcomes.

#### Subgroup Analysis

To address possible effect modification, we will use the same analytical approach as specified for the primary and secondary outcomes but include a treatment by subgroup interaction term in the model. Previous research found presence of ureteral stent(s) as a potential risk factor for UTI in patients undergoing SWL [4]. We will therefore explore how ciprofloxacin versus placebo impacts patients with and without ureteral stents. We expect ciprofloxacin to have larger relative (and absolute) benefit in patients having ureteral stents. We will report results using a forest plot illustrating risk ratios with 95% confidence intervals and in accordance with best practices and guidelines for subgroup analyses [22–24]. We will use the Instrument for assessing the Credibility of Effect Modification Analyses (ICEMAN) to guide inferences about the credibility of our subgroup analyses [22].

#### Sensitivity Analyses

If the primary analysis yields a significant result for the primary outcome, we will conduct sensitivity analyses to assess the robustness of the findings regarding the impact of missing data [25,26]. We will assume that participants in the placebo group (control arm) with missing data have the same event rate as those who completed follow-up in the placebo group.

We will first assume that participants in the ciprofloxacin group (intervention arm) with missing data have double the event rate of those who completed follow-up in the ciprofloxacin group. If the result remains significant, we will proceed stepwise, increasing the assumed event rate to triple, then four times, and finally five times – but only if each preceding analysis remains significant. If an assumption fails to yield a significant result, we will not proceed further.

#### Interim Analysis

A blinded (pooled data from both treatment groups) re-estimation of the sample size will be conducted when at approximately 66% (n=1000) of patients have been recruited if the observed event rate is lower than the expected event rate.

### Dissemination of the results

After the end of the study, the coordinating centre undertakes to publish the results, remaining the sole owner of the data. All publications will undergo a review by the Steering Committee before submission. We will present the trial results according to the Consolidated Standards of Reporting Trials (CONSORT) guidelines for RCTs [27]. The results of the APPEAL will provide the highest quality evidence available to inform future clinical practice. APPEAL also created a large trial network that will facilitate future studies and therefore represented a crucial step to establish CLUE Working Group as trial unit of excellence focused on global, pragmatic RCTs [28].

## Discussion

Large discrepancies exist between guidelines and clinical practice patterns regarding antibiotic prophylaxis for patients undergoing SWL. Given the low frequency of severe infectious complications following SWL, a large pragmatic RCT is necessary to determine the need for prophylaxis and identify the specific patient populations that may benefit.

This trial commenced recruitment in London (Ontario, Canada) in 2013 as a single-centre trial. Recruitment challenges are common in randomized surgical trials: slow recruitment is the most common reason for discontinuation [29]. This trial faced similar challenges. Owing to slow recruitment, we expanded the trial globally and renamed as APPEAL.

In 2019, the European Commission issued a legally binding decision restricting the use of quinolone antibiotics, including ciprofloxacin, across all EU countries, primarily because of adverse effects associated with prolonged use [30]. This decision has influenced opinions on suitable antibiotic prophylaxis strategies, particularly in the context of urinary procedures for which quinolones were commonly used. Despite this, we believed that continuation of recruitment for APPEAL was ethical because i) evidence has shown that the adverse effects of quinolones are overwhelmingly associated with long-term use, and ii) ciprofloxacin remains one of the prophylactic antibiotics most often used in urology.

The results of our RCT, whether positive or negative, will significantly contribute to the existing literature by providing high-certainty evidence with the largest patient population to date for future meta-analyses and guideline development. If the study demonstrates a benefit from antibiotic use, it will reduce the rates of infectious complications in patient care. Additionally, our study may enable a more selective use of antibiotics in line with recent guidelines by identifying key predictors of infection. Conversely, a negative result will offer the strongest evidence to date against routine prophylaxis, potentially decreasing the unnecessary use of antibiotics among SWL patients. This could lead to cost savings for the healthcare system and reduce the risk of antibiotic-resistant bacteria. Furthermore, we propose evaluating a straightforward antibiotic protocol that any institution can easily adopt. Therefore, the results of this study will have a significant impact on recommendations for the use of pre-SWL antibiotics.

## Data Availability

All data produced in the present work are contained in the manuscript.

---

Appendix 1. Details on our survey methods and results

Appendix 2. Ethics decisions/approvals of the local institutional review boards

Appendix 3. A modified IPSS questionnaire

## References

1. Becerra AZ, Khusid JA, Sturgis MR et al. Contemporary Assessment of the Economic Burden of Upper Urinary Tract Stone Disease in the United States: Analysis of One-Year Health Care Costs, 2011– 2018. J Endourol. 2021;36(4):429–38.

2. Lang J, Narendrula A, El-Zawahry A, Sindhwani P, Ekwenna O. Global Trends in Incidence and Burden of Urolithiasis from 1990 to 2019: An Analysis of Global Burden of Disease Study Data. Eur Urol Open Sci. 2022;35:37–46.

3. Setthawong V, Srisubat A, Potisat S, Lojanapiwat B, Pattanittum P. Extracorporeal shock wave lithotripsy (ESWL) versus percutaneous nephrolithotomy (PCNL) or retrograde intrarenal surgery (RIRS) for kidney stones. Cochrane Database Syst Rev. 2023;8(8):CD007044.

4. Duvdevani M, Lorber G, Gofrit ON, Latke A, Katz R, Landau EH, et al. Fever after shockwave lithotripsy--risk factors and indications for prophylactic antimicrobial treatment. J Endourol. 2010;24(2):277–81.

5. Shafi H, Ilkhani M, Darabi Ahangar Z, Bayani M. Antibiotic prophylaxis in the prevention of urinary tract infection in patients with sterile urine before extracorporeal shock wave lithotripsy. Caspian J Intern Med. 2018;9(3):296–8.

6. Na L, Li J, Pan C, Zhan Y, Bai S. Development and validation of a predictive model for major complications after extracorporeal shockwave lithotripsy in patients with ureteral stones: based on a large prospective cohort. Urolithiasis. 2023;51(1):42.

7. Alexander CE, Gowland S, Cadwallader J, Hopkins D, Reynard JM, Turney BW. Routine Antibiotic Prophylaxis Is Not Required for Patients Undergoing Shockwave Lithotripsy: Outcomes from a National Shockwave Lithotripsy Database in New Zealand. J Endourol. 2016;30(11):1233–8.

8. Mira Moreno A, Montoya Lirola MD, García Tabar PJ, Galiano Baena JF, Tenza Tenza JA, Lobato Encinas JJ. Incidence of Infectious Complications after Extracorporeal Shock Wave Lithotripsy in Patients Without Associated Risk Factors. J Urol. 2014;192(5):1446–9.

9. Lantz AG, McKay J, Ordon M, Pace KT, Monga M, Honey RJDA. Shockwave Lithotripsy Practice Pattern Variations Among and Between American and Canadian Urologists: In Support of Guidelines. J Endourol. 2016;30(8):918–22.

10. Doherty R, Manley K, Gordon S et al. Current ESWL practice and outcomes in the UK: A multicentre snapshot. J Clin Urol. 2017;10(4):340–6.

11. Bonkat G, Bartoletti R, Bruyère F et al. EAU Guidelines on Urological Infections. Arnhem, the Netherlands: EAU Guidelines Office. 2024.

12. Skolarikos A, Jung H, Neisius A et al. EAU Guidelines on Urolithiasis. Arnhem, the Netherlands: EAU Guidelines Office. 2024.

13. Mrkobrada M, Ying I, Mokrycke S et al. CUA Guidelines on antibiotic prophylaxis for urologic procedures. Can Urol Assoc J. 2015;9(1-2):13–22.

14. Lightner DJ, Wymer K, Sanchez J, Kavoussi L. Best Practice Statement on Urologic Procedures and Antimicrobial Prophylaxis. J Urol. 2020;203(2):351–6.

15. Lu Y, Tianyong F, Ping H, Liangren L, Haichao Y, Qiang W. Antibiotic prophylaxis for shock wave lithotripsy in patients with sterile urine before treatment may be unnecessary: a systematic review and meta-analysis. J Urol. 2012;188(2):441–8.

16. Assimos D, Krambeck A, Miller NL et al. Surgical Management of Stones: American Urological Association/Endourological Society Guideline, PART I. J Urol. 2016;196(4):1153–60.

17. Memmos D, Mykoniatis I, Sountoulides P et al. Evaluating the usefulness of antibiotic prophylaxis prior to ESWL in patients with sterile urine: a systematic review and meta-analysis. Minerva Urol Nephrol. 2021;73(4):452–61.

18. Tikkinen KAO, Tornberg SV, Ruotsalainen J et al. Update on APPEAL, an International Randomized Controlled Trial Evaluating Ciprofloxacin Versus Placebo in Patients Undergoing Shockwave Lithotripsy for Urolithiasis. Eur Urol Focus. 2024;10(5):697–9.

19. Wolf JS Jr, Bennett CJ, Dmochowski RR, Hollenbeck BK, Pearle MS, Schaeffer AJ. Best practice policy statement on urologic surgery antimicrobial prophylaxis. J Urol. 2008;179(4):1379–90.

20. Chen W, Qian L, Shi J, Franklin M. Comparing performance between log-binomial and robust Poisson regression models for estimating risk ratios under model misspecification. BMC Med Res Methodol. 2018;18(1):63.

21. Zou G. A Modified Poisson Regression Approach to Prospective Studies with Binary Data. Am J Epidemiol. 2004;159(7):702–6.

22. Schandelmaier S, Briel M, Varadhan R et al. Development of the Instrument to assess the Credibility of Effect Modification Analyses (ICEMAN) in randomized controlled trials and meta-analyses. CMAJ. 2020;192(32):E901–e6.

23. Sun X, Briel M, Walter SD, Guyatt GH. Is a subgroup effect believable? Updating criteria to evaluate the credibility of subgroup analyses. BMJ. 2010;340:c117.

24. Sun X, Ioannidis JP, Agoritsas T, Alba AC, Guyatt G. How to use a subgroup analysis: users’ guide to the medical literature. JAMA. 2014;311(4):405–11.

25. Akl EA, Briel M, You JJ et al. Potential impact on estimated treatment effects of information lost to follow-up in randomised controlled trials (LOST-IT): systematic review. BMJ. 2012;344:e2809.

26. Akl EA, Johnston BC, Alonso-Coello P et al. Addressing dichotomous data for participants excluded from trial analysis: a guide for systematic reviewers. PLoS One. 2013;8(2):e57132.

27. Turner L, Shamseer L, Altman DG et al. Consolidated standards of reporting trials (CONSORT) and the completeness of reporting of randomised controlled trials (RCTs) published in medical journals. Cochrane Database Syst Rev. 2012;11(11):Mr000030.

28. Ng A, Asif A, Keane K, Ippoliti S, Nathan A, Kasivisvanathan V. The ARTS (Avoiding Risks of Thrombosis and Bleeding in Surgery) Trial: Lessons Learnt in Setting Up an International Multicentre Clinical Trial of an Investigational Medicinal Product in the UK. Eur Urol Focus. 2023;9(5):695–7.

29. Rosenthal R, Kasenda B, Dell-Kuster S et al. Completion and Publication Rates of Randomized Controlled Trials in Surgery: An Empirical Study. Ann Surg. 2015;262(1):68–73.

30. European Medicines Agency. Quinolone- and fluoroquinolon econtaining medicinal products. Amsterdam, The Netherlands: European Medicines Agency; 2019. https://www.ema.europa.eu/en/medicines/human/referrals/quinolone-fluoroquinolone-containing-medicinal-products [

