## Appendices 1-3 for "A Multicentre Randomised Controlled Trial Assessing the Efficacy of Antimicrobial Prophylaxis for Extracorporeal Shock Wave Lithotripsy in Reducing Urinary Tract Infection (APPEAL): Statistical Analysis Plan and Methodology"

**Appendix 1.** Details on our survey methods and results

**Methods:** We investigated potential variations in the use of antibiotic prophylaxis before or after shock wave lithotripsy by surveying the lead investigators at each APPEAL trial site. The survey focused on their clinical practices prior to the initiation of the APPEAL trial at their respective sites, using the questions provided in the supplementary table 1 below.

**Supplementary Table 1**. Typical use of antibiotic prophylaxis for shock wave lithotripsy (SWL) before the APPEAL trial.

CENTER NAME:
PERSON WHO RESPONDED:
DATE:

| **Survey Questions** | **For patients without ureteral (JJ) stent:** | **For patients WITH ureteral (JJ) stent:** |
|---|---|---|
| 1. Did you typically use antibiotics (before/outside the APPEAL trial)? | 0. No<br>1. Yes | 0. No<br>1. Yes |
| 2a. **If you answered yes**, which antibiotic: | 1. Ciprofloxacin<br>2. Sulfamethoxazole<br>3. Co-amoxiclav<br>4. Other (describe) | 1. Ciprofloxacin<br>2. Sulfamethoxazole<br>3. Co-amoxiclav<br>4. Other (describe) |
| 2b. **If you answered yes**, what is the duration of antibiotic treatment? | 1. Single dose before SWL<br>2. One dose pre-SWL and one dose post-SWL<br>3. Other (describe) | 1. Single dose before SWL<br>2. One dose pre-SWL and one dose post-SWL<br>3. Other (describe) |
| 2c. **If you answered yes**, how antibiotics have been given? | 1. Oral<br>2. Intravenous<br>3. Other (describe) | 1. Oral<br>2. Intravenous<br>3. Other (describe) |

**Results:** Out of the 12 centers, 5 centers (42%) prescribed antibiotic prophylaxis for patients without a stent, 9 centers (75%) did so for patients with a stent, and 3 centers (25%) never prescribed antibiotic peri-SWL. A detailed description of these results is provided in the supplementary table 2 below. Although the sample size was small, the results reveal no single dominant approach to antibiotic prophylaxis use prior to SWL, regardless of whether a JJ stent was employed.

**Supplementary Table 2.** Current practices of antibiotic prophylaxis use prior to shockwave lithotripsy at study sites participating in the APPEAL trial. Responses sorted according to readiness to provide antibiotic prophylaxis as standard practice prior to shockwave lithotripsy.

| Study site | Typical use of antibiotic prophylaxis for SWL before APPEAL trial | | | | If yes, which antibiotic(s) were used? |
|---|---|---|---|---|---|
| | Without JJ stent | | With JJ stent | | |
| | Yes | Not typically | Yes | Not typically | |
| **Prescribed in both** | | | | | |
| Jeddah (SA) | ● | | ● | | Ciprofloxacin |
| Moscow (RUS) | ● | | ● | | Cephalosporin 1.0 I.M. BID, starting pre-SWL, for 2-3 days |
| Sherbrooke (CAN) | ● | | ● | | Ciprofloxacin |
| St. Petersburg 2 (RUS) | ● | | ● | | Single dose of ceftriaxone |
| Tabriz (IR) | ● | | ● | | Ciprofloxacin |
| **Prescribed only in patients with stent** | | | | | |
| Basel (CH) | | ● | ● | | Sulfamethoxazole/Trimethoprim |
| Helsinki (FIN) | | ● | ● | | Ciprofloxacin |
| London (CAN) | | ● | ● | | Ciprofloxacin or cefazolin (I.V.) |
| Sao Paulo (BR) | | ● | ● | | Ciprofloxacin |
| **Never prescribed** | | | | | |
| Jakarta (ID) | | ● | | ● | |
| Doha (QA)* | | ● | | ● | |
| St. Petersburg 1 (RUS) | | ● | | ● | |

* In Doha, shock wave lithotripsy is limited to patients with a low white blood cell count and a negative urine culture.

**Appendix 2.** Ethics decisions/approvals of the local institutional review boards

| Center | Decision number | Ethics Committee name |
|---|---|---|
| Basel | 2019–01425 | Ethikkommission Nordwest- und Zentralschweitz (EKNZ) |
| Doha | MRC-01-21-295 | Institutional Review Board Hamad Medical Corporation |
| Helsinki | 155/13/03/02/16 | Helsingin ja Uudenmaan sairaanhoitopiiri Operatiivinen eettinen toimikunta |
| Jakarta | KET-472/UN2.F1/ ETIK/PPM.00.02/2021 | Komite Etik Penelitian Kesehatan Fakultas Kedokteran Universitas Indonesia - RSUPN |
| Jeddah | 555–17 | King Abdulaziz University, Faculty of Medicine Unit of Biomedical Ethics Research Committee |
| London (Ontario) | REB 103696 | The University of Western Ontario Health Sciences Research Ethics Board (HSREB) |
| Moscow | 23-21_17.12.2021 | Sechenov University IRB |
| Sao Paolo | 2648767 | Ethics Committee of the Hospital e Maternidade São Luis |
| Sherbrooke | #2017-1639 | Research Ethics Board of the Centre intégré universitaire de santé et de services sociaux de l'Estrie - Centre hospitalier universitaire de Sherbrooke (CIUSSSE-CHUS) |
| St. Petersburg | #219 (27/05/2019) | Ethics Committee of Saint-Petersburg State Medical University |
| Tabriz | IR.TBZMED.REC.1400.131 | Tabriz University of Medical Sciences (Biomedical Research Ethics Committee) |

**Appendix 3.** A modified IPSS questionnaire

Study ID: _____________

Pre-SWL ☐ / Post-SWL: ☐ Date: ___/______/___
(day / month / year)

Please complete the following questionnaire **by circling the most appropriate response:**

| | Not at all | Less than 1 time in 5 | Less than half the time | About half the time | More than half the time | Almost always |
|---|---|---|---|---|---|---|
| Incomplete emptying<br>In the past week, how often have you had a sensation of not emptying your bladder completely after you finish urinating? | 0 | 1 | 2 | 3 | 4 | 5 |
| Frequency<br>In the past week, how often have you had to urinate again less than two hours after you finished urinating? | 0 | 1 | 2 | 3 | 4 | 5 |
| Intermittency<br>In the past week, how often have you found you stopped and started again several times when you urinated? | 0 | 1 | 2 | 3 | 4 | 5 |
| Urgency<br>In the past week, how difficult have you found it to postpone urination? | 0 | 1 | 2 | 3 | 4 | 5 |
| Weak stream<br>In the past week, how often have you had a weak urinary stream? | 0 | 1 | 2 | 3 | 4 | 5 |
| Straining<br>In the past week, how often have you had to push or strain to begin urination? | 0 | 1 | 2 | 3 | 4 | 5 |
| Pain<br>In the past week, how often have you had burning or discomfort with urination? | 0 | 1 | 2 | 3 | 4 | 5 |

| | None | 1 time | 2 times | 3 times | 4 times | 5 times or more |
|---|---|---|---|---|---|---|
| Nocturia<br>In the past week, how many times did you most typically get up to urinate from the time you went to bed until the time you got up in the morning? | 0 | 1 | 2 | 3 | 4 | 5 |

| | Delighted | Pleased | Mostly satisfied | Mixed – about equally satisfied | Mostly dissatisfied | Unhappy | Terrible |
|---|---|---|---|---|---|---|---|
| If you were to spend the rest of your life with your urinary condition the way it is now, how would you feel about that? | 0 | 1 | 2 | 3 | 4 | 5 | 6 |
